# Prevalence of Family Planning Utilization and Its Determinants among Women of Reproductive Age in Oromia and Gambella Regions, Ethiopia

**DOI:** 10.64898/2026.03.06.26347771

**Authors:** Kunuz Hajibedru Abadula, Abebaw Gebeyehu Worku, Gurmesa Tura Debelew, Muluemebet Abera Wordofa

## Abstract

**Background:** Family planning (FP) is essential for improving maternal and child health. Despite Ethiopia’s national progress, regional disparities persist in underserved areas. This study assesses FP utilization and its determinants among women of reproductive age in the Oromia and Gambella regions.

**Methods:** A community-based cross-sectional study was conducted from October 15–25, 2023, among 840 women of reproductive age selected from five districts in Oromia and Gambella Regions. Data were collected using a structured, interviewer-administered questionnaire adapted from the Demographic and Health Survey and implemented through SurveyCTO. Multivariable logistic regression analysis was performed to identify factors associated with FP utilization, with adjusted odds ratios (AORs) and 95% confidence intervals (CIs) reported.

**Results:** FP utilization was 60.9%, with injectables (48.2%) and implants (30.4%) being most common. Utilization was significantly lower among women lacking transport access (AOR=0.49, 95% CI: 0.36–0.67) and those in the poorest (AOR=0.48, 95% CI: 0.29–0.77) and poor (AOR=0.47, 95% CI: 0.29–0.74) wealth quintiles. Women whose partners had no formal education (AOR=0.46, 95% CI: 0.30–0.70) or only primary education (AOR=0.64, 95% CI: 0.44–0.92) were less likely to use FP compared to those with more educated partners. Additionally, farming women were more likely to use FP (AOR=1.64, 95% CI: 1.11–2.42), while those reporting unwanted pregnancies had lower utilization (AOR=0.54, 95% CI: 0.32–0.92).

**Conclusion:** FP utilization in these regions exceeds national averages, yet reliance on short-acting methods remains high. Limited transport, low household wealth, low partner education, and pregnancy unintendedness are critical barriers. Strengthening community-based services, addressing economic disparities, and promoting male involvement are essential for improving equitable FP access.

## Background

Family planning (FP) is vital for reducing maternal and child morbidity and mortality and promoting gender equity and development (1–3). Despite its benefits, unmet FP needs remain high in low- and middle-income countries, especially sub-Saharan Africa, where 270 million women lacked access to modern contraception in 2021, contributing to millions of unintended pregnancies, unsafe abortions, and maternal deaths annually (4–6). Between 2012 and 2025, modern contraceptive use doubled in 28 FP2030 countries, and unmet need in sub-Saharan Africa declined from 21.1% to 18.5% (7). FP uptake is influenced by education, wealth, employment, media exposure, antenatal care, and family size, and lower among younger women, those with fewer health visits, or prior abortion (8,9). Country-specific evidence shows that women’s empowerment, spousal communication, and community education reduce unmet need, while cultural norms, rural residence, and absent partners limit use (10–12).

In sub-Saharan Africa, married women’s decision-making power and use of long-acting contraceptives are positively associated with education, wealth, employment, media exposure, antenatal care, and larger family size, but lower among younger women, those with fewer health visits, delayed sexual debut, or prior abortion, highlighting key socio-economic, reproductive, and informational determinants (8,9). FP uptake is shaped by broader socio-cultural and gender factors (8–10). Country-specific evidence shows that women’s empowerment, prior contraceptive use, and community education reduce unmet need in Tanzania (11); education, economic empowerment, spousal communication, and autonomy influence FP choices in Ghana, Kenya, and Nigeria (9); and in Cameroon, modern contraceptive use is higher in wealthier, smaller households with educated husbands but lower where husbands are absent or cultural norms favor multiple male children (12).

In Ethiopia, FP has been prioritized through the Health Extension Program and SDG commitments, increasing contraceptive prevalence from 8% in 2000 to 41.4% in 2019 and reducing unmet need from 37% to 22% (13–16). Despite these gains, universal and equitable coverage is hindered by resource shortages, weak governance, rural inequities, and gaps in infrastructure, workforce, and logistics. Regional and socio-demographic disparities persist, with higher FP use in Oromia and Amhara than Somali, influenced by residence, education, marital status, media exposure, and professional counseling, while unmet need is spatially clustered along urban–rural lines (17–19).

In Oromia, many women switch from long-acting to short-acting contraceptives due to fear of side effects, misconceptions, and limited information, while marginalized women face barriers related to age, income, and living conditions (20,21). National modern contraceptive use is 37–38%, higher among younger, educated, urban, and wealthier women, with limited decision-making autonomy (22,23). In Gambella, prevalence is 25.5%, influenced by age, religion, education, marital status, family size, knowledge, attitudes, and sociocultural factors (24). This study examines socioeconomic, demographic, and access-related determinants of FP utilization in Ethiopia to inform policies for universal coverage.

## Methods

### Setting Area

This study was conducted in Health Information System (HIS) intervention sites supported by Jimma University, one of six institutions selected to implement the CBMP interventions. The intervention was initially rolled out across seven zones in Oromia Region and two zones in Gambella Region. Data were collected from five woredas—Omo Nada, Metu Zuria, Boset, and Digalu/Tijo in Oromia, and Gambella Zuria in Gambella. These study areas have a combined population of 775,078 and are served by 26 health centers and 124 health posts, representing diverse geographic settings and health system contexts.

### Study design

This community-based cross-sectional study was conducted to assess the utilization of FP services and its determinants across populations with varying socioeconomic and demographic characteristics. The design was selected for its efficiency in generating preliminary evidence on FP use at a single point in time. Data collection was carried out from **October 15 to 25, 2023**.

### Ethical Considerations

Ethical clearance for the study was obtained from the **Jimma University Institute of Health Institutional Review Board (IRB)** (Ref. No: JUIH/IRB/586/23, dated August 23, 2023). Written informed consent was obtained from all participants prior to data collection. Confidentiality was maintained by coding responses and ensuring no personally identifiable information was accessible to researchers.

### Study Population and Eligibility Criteria

The study population consisted of women of reproductive age residing in the catchment areas of selected health facilities. A total of 840 women were recruited from 12 health centers serving 24 kebeles. Eligibility was restricted to women who were permanent residents in the study area and within the reproductive age group during the data collection period. Women who were critically ill at the time of the survey or who declined to participate were excluded from the study.

### Sample size and sampling techniques

The sample size was determined using the single population proportion formula, assuming a modern contraceptive prevalence of 38% based on national survey data (25). With a 95% confidence level and 5% margin of error, the initial calculation yielded 384 participants. After applying a design effect of 2 to account for clustering and adding 10% for potential non-response, the final sample size was 840 women of reproductive age.

A multistage sampling approach was applied: 12 health centers were purposively selected, two kebeles randomly chosen per facility, and 840 eligible women systematically drawn from household lists across 24 kebeles using health post records or a pre-survey census The household survey sampling frame was developed using either health post records or a pre-survey census, depending on data availability. Each kebele, Ethiopia’s smallest administrative unit with about 1,000 households, offered a reliable basis for community-level sampling and ensured broad coverage of the target population.

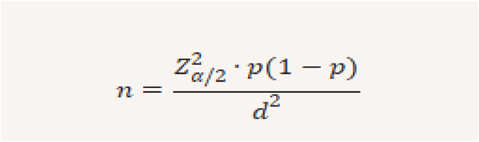

### Data Collection Procedures and Tool

Data collection was carried out using a structured questionnaire adapted from the Demographic and Health Survey (DHS) and administered through the SurveyCTO digital platform. The instrument was translated into **Oromiffa, Amharic, and Anuak** to ensure local understanding. A team of **24 enumerators and five supervisors** received three days of training covering study protocols, ethical standards, and use of the electronic system. Responses were captured on mobile devices, allowing for real-time monitoring and quality control. Prior to the main survey, the questionnaire was pilot-tested to confirm clarity and consistency.

## Variables

### Measurement of the Primary Outcome

The primary outcome of this study was FP utilization, categorized as *users* (women currently using any contraceptive method) and *non-users* (women not using contraception at the time of the survey). Data were collected through self-report and coded as a binary variable (“1” for users, “0” for non-users). For analysis, this dichotomous outcome was applied directly in logistic regression models to examine associations with socioeconomic and demographic determinants.

### Independent variables

The independent variables examined in relation to FP utilization included socio-demographic factors (school attendance, mother’s educational status, partner’s educational status), economic factors (household wealth index categories, mother’s occupation, mother’s employment status), reproductive factors (pregnancy wontedness), and access-related factors (availability of transport and proximity to health facilities).

### Data Processing and Statistical Analysis

Data were collected electronically through SurveyCTO, exported, and checked for accuracy before analysis in **SPSS version 25**. Descriptive statistics were used to summarize participant characteristics. Associations with FP utilization were first explored using bivariable logistic regression; variables with a p-value below 0.25 were then entered into a multivariable model. A backward stepwise procedure was applied to control for potential confounders. Final results are presented as adjusted odds ratios (AORs) with 95% confidence intervals, and statistical significance was set at p < 0.05.

Data were collected electronically using SurveyCTO, then downloaded, checked for completeness, and cleaned before analysis in SPSS Version 25.0. Descriptive statistics summarized participant characteristics. To identify factors associated with maternal CoC completion, a bivariable analysis was conducted; all variables with a p-value < 0.25 were included in a subsequent multivariable logistic regression model. A backward stepwise approach was used to build the final model, controlling for confounders. Results are reported as adjusted odds ratios (AORs) with 95% confidence intervals, with statistical significance set at p < 0.05. Multicollinearity was assessed using variance inflation factors (VIF) and tolerance values in SPSS version 25; all VIFs were below 10 and tolerance values above 0.10, indicating multicollinearity was not a concern.

## Results

### Descriptive Summary of Respondents’ Characteristics

A total of 840 women of reproductive age participated in the study, with the majority (51.7%) aged 25–34 years, followed by 38.5% in the 15–24 age groups. Most participants were married 98%. Regarding educational attainment, 71.2% of mothers had attended school, with 50.4% completing primary education. Partners’ education followed a similar pattern, with 45.2% having completed primary school and 31.8% attaining secondary education.

The majority of mothers (92.4%) were unemployed or engaged in subsistence farming. Similarly, the majority of partners were engaged in farming (88.2%), while only 11.8% were employed in formal occupations. Wealth distribution was relatively balanced across quintiles, with 18.8% classified as poorest, 21.1% and 21.2% richest. Regarding pregnancy wontedness, 90.7% of respondents reported their most recent pregnancy was wanted.

Overall, these descriptive statistics indicate that the study population was predominantly young, married, and farmers, with low formal employment, moderate levels of educational attainment, and mostly wanted pregnancies. These background characteristics provide important context for interpreting patterns of FP utilization in Oromia and Gambella regions Table 1.

**Table 1:**
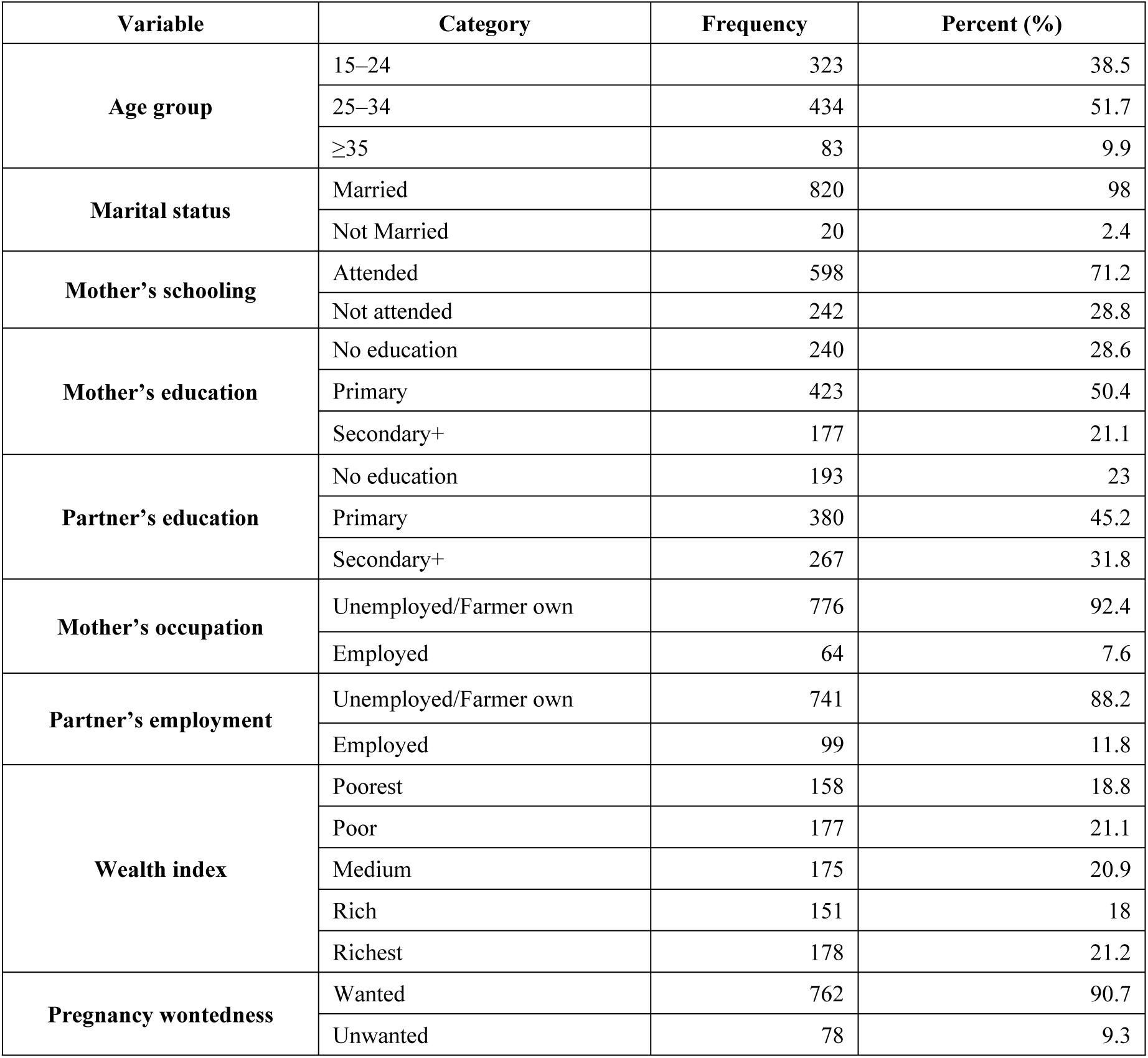
Background Characteristics of Participants: Family Planning Utilization in Oromia and Gambella, Ethiopia, October 2023.

### Knowledge of FP Methods

Out of 840 respondents, 651 (77.5%) provided valid responses regarding knowledge of FP methods. Awareness was highest for modern methods, with injectable (39.0%) and implants (29.5%) most frequently mentioned. Pills (12.4%) and condoms (5.5%) were moderately known, while knowledge of intrauterine devices (3.8%), lactation amenorrhea (4.6%), and rhythm (1.2%) was limited. Sterilization, female condoms, and emergency contraception were rarely reported (<1%). These findings indicate that while knowledge of modern methods is widespread, awareness of long-acting reversible and traditional methods remains relatively low Figure 1

**Figure 1.**
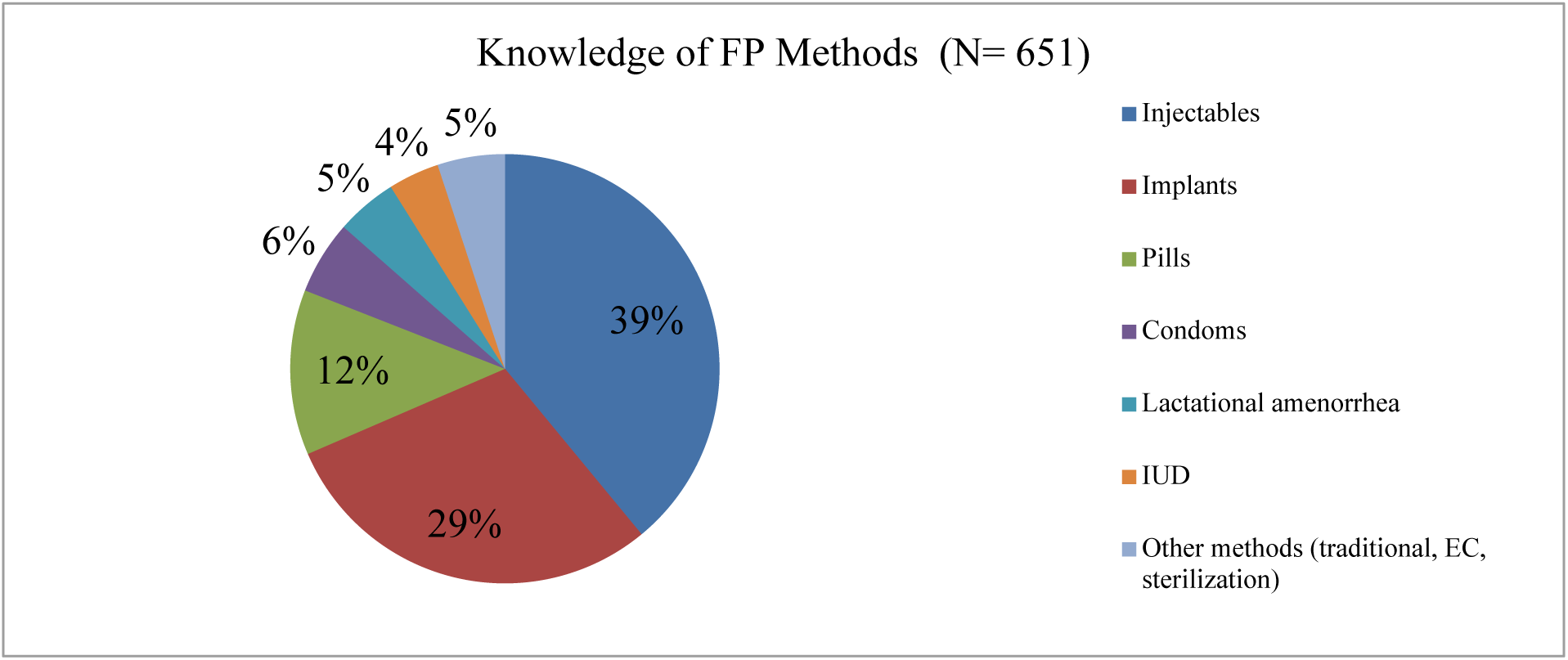
Knowledge of Family Planning Methods among Respondents (N = 651)

### Main Methods Used

Descriptive analysis showed that 60.9% of women currently used family planning, with modern methods predominating. Injectables were the most common method (48.2%), followed by implants (30.4%). Pills, lactational amenorrhea, and other methods such as condoms, IUDs, and traditional practices each accounted for about 7%. This distribution highlights a strong reliance on short-acting methods, with injectables and implants together representing nearly 80% of contraceptive use.

**Figure 2.**
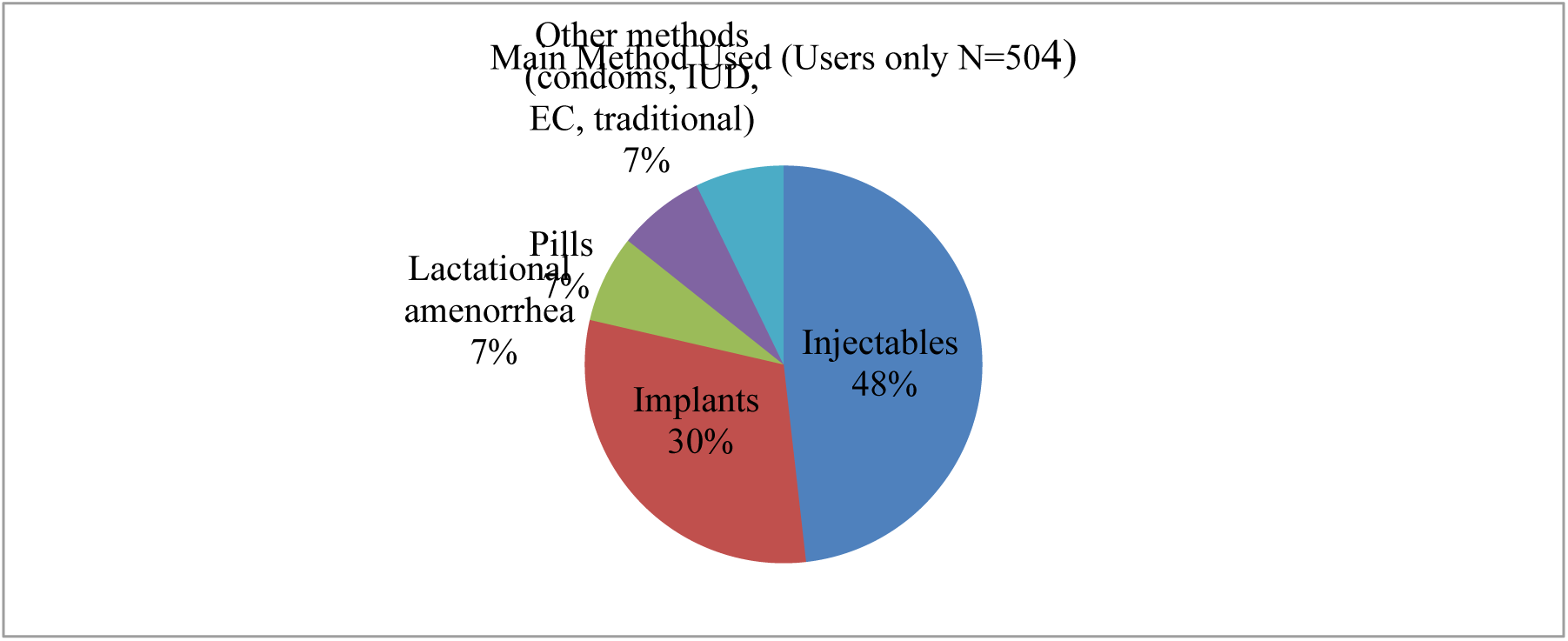
Main Methods of Family Planning Used among Respondents (N = 504)

### Method Type

Overall, 68.8% of women relied on short-acting methods, compared to 31.2% who used long-acting options. The predominance of short-acting methods reflects limited uptake of long-term contraceptives, despite their effectiveness and convenience. This imbalance underscores the need for improved counseling and access to long-acting reversible contraceptives to support sustained reproductive planning.

**Figure 3.**
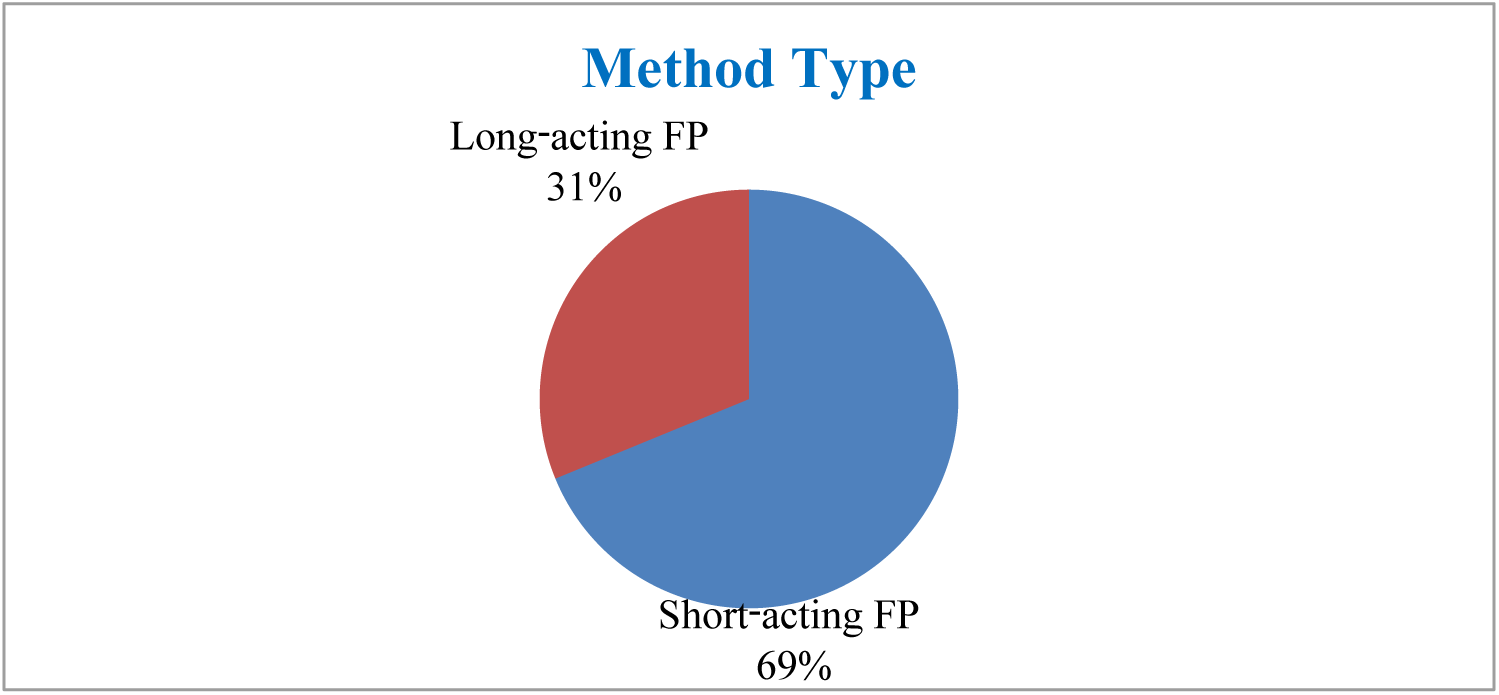
Method Type of Family Planning Used among Respondents (N = 504)

### Source of Family Planning Methods

Among 501 respondents reporting their contraceptive sources, government health centers (51.5%) and health posts (32.7%) provided over 84% of FP services, while government hospitals (1.8%) and private clinics (1.4%) contributed minimally. Home-based sources accounted for 12.6%, reflecting the continued role of informal or self-administered channels, particularly for traditional methods and self-care, and highlighting the central role of Ethiopia’s public health system in modern contraceptive provision **Table 2**

**Table 2.**
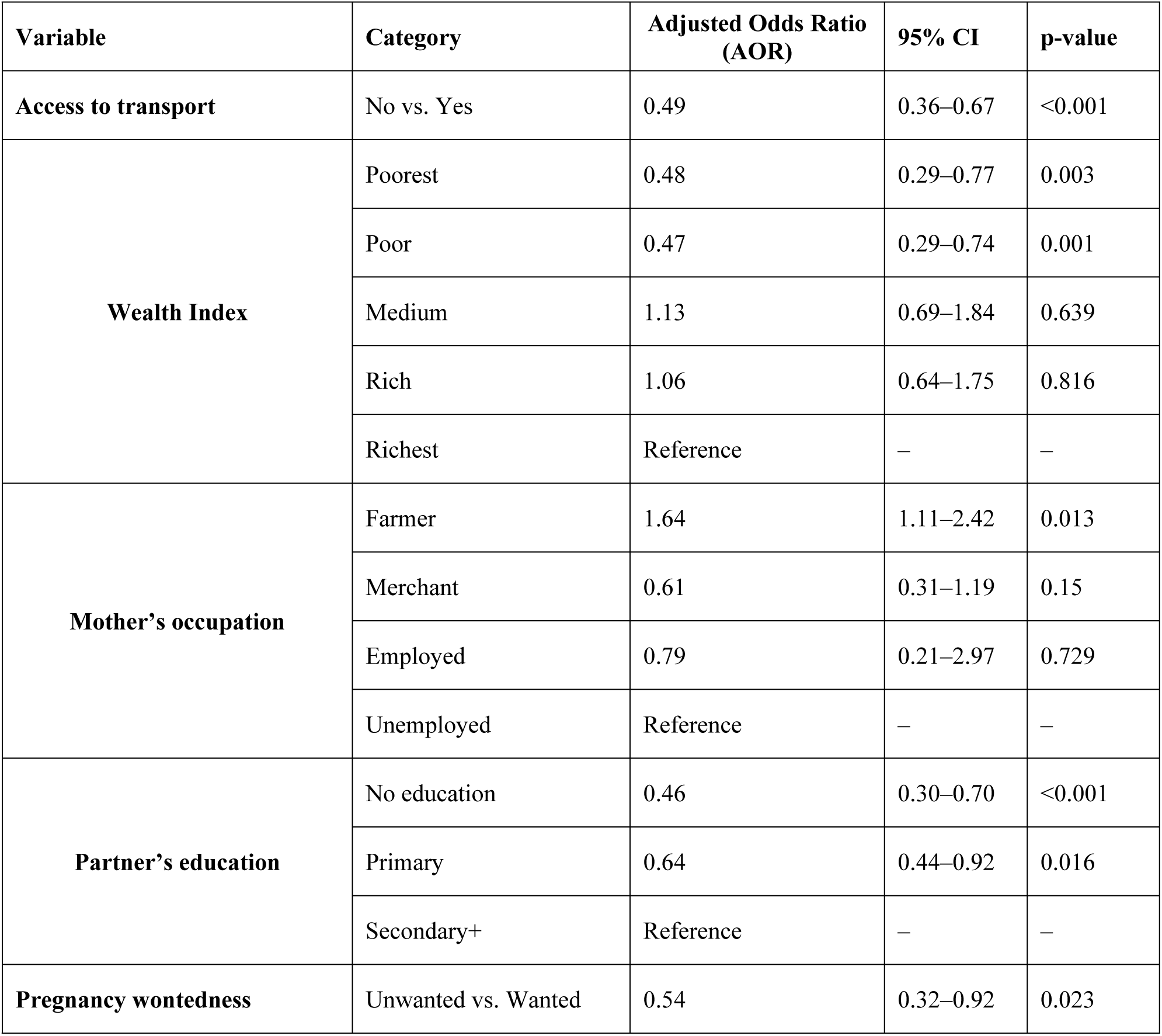
Determinants of Family Planning Utilization among Women of Reproductive Age in Oromia and Gambella, Ethiopia (N = 840)

**Figure 4.**
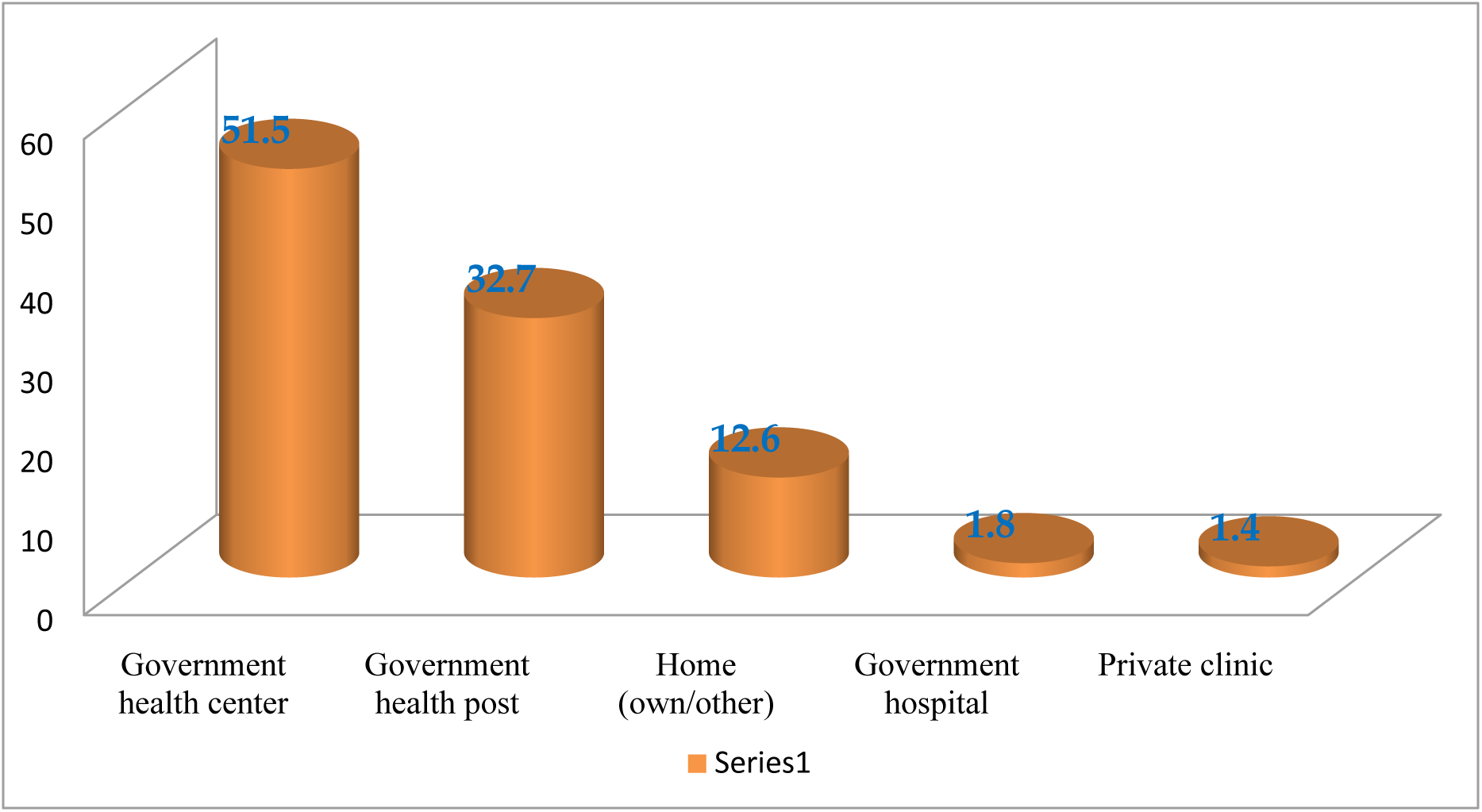
Source of Family Planning among Respondents (N = 504)

### Factors Associated with Determinants of Family Planning Utilization

Multivariable logistic regression analysis was conducted to identify factors associated with family planning utilization among 840 women of reproductive age in Oromia and Gambella regions. The results indicate that both individual and household-level characteristics significantly influenced FP use.

Women who lacked access to transportation were significantly less likely to use family planning compared with those who had access (AOR = 0.49, 95% CI: 0.36–0.67, p < 0.001). Household wealth was also an important determinant: women in the poorest (AOR = 0.48, 95% CI: 0.29–0.77, p = 0.003) and poor (AOR = 0.47, 95% CI: 0.29–0.74, p = 0.001) wealth categories were significantly less likely to use FP compared with women in the richest category, whereas those in the medium (AOR = 1.13, 95% CI: 0.69–1.84, p = 0.639) and rich (AOR = 1.06, 95% CI: 0.64–1.75, p = 0.816) categories did not differ significantly.

Maternal occupation also influenced contraceptive use. Women engaged in farming were more likely to use FP than unemployed women (AOR = 1.64, 95% CI: 1.11–2.42, p = 0.013). while being a merchant (AOR = 0.61, 95% CI: 0.31–1.19, p = 0.150) or formally employed (AOR = 0.79, 95% CI: 0.21–2.97, p = 0.729) was not significantly associated with FP utilization.

Partner’s educational status was positively associated with women’s FP use. Women whose partners had no formal education were less likely to use FP (AOR = 0.46, 95% CI: 0.30–0.70, p < 0.001), and those whose partners had primary education were also less likely (AOR = 0.64, 95% CI: 0.44–0.92, p = 0.016) compared with women whose partners had secondary education or higher.

Finally, pregnancy wontedness was a significant predictor: women reporting an unwanted pregnancy were less likely to use FP than those with wanted pregnancies (AOR = 0.54, 95% CI: 0.32–0.92, p = 0.023).

Overall, these findings highlight that limited access to transport, low household wealth, low partner education, maternal occupation, and pregnancy intenseness are important determinants of family planning utilization in these regions. Interventions targeting these factors may improve equitable access to FP services **Table 2**.

## Discussion

Our findings reflect the demographic profile of rural Ethiopian women, where fertility norms and limited autonomy shape contraceptive behavior (25,26). Despite high awareness, knowledge remains skewed toward short-acting methods particularly injectables and implants while awareness of long-acting options is limited, restricting informed choice (32,33). The relatively higher uptake of long-acting methods observed in this study (31.2%) exceeds national averages and may be linked to strengthened Health Information System (HIS) interventions, which improve service monitoring, commodity forecasting, and accountability (40,41) (42,43). Service delivery remains concentrated in government health centers and health posts, with minimal contributions from hospitals and private clinics, underscoring the central role of Ethiopia’s public health system and the need to expand community-based distribution and outreach (16).

Several interrelated socioeconomic and interpersonal factors further influenced FP uptake. Women without reliable transport were substantially less likely to use FP, highlighting persistent geographic barriers in rural Ethiopia (44,45). Wealth status showed a clear gradient: women in lower wealth quintiles were less likely to use FP, reflecting how poverty constrains access to information, transportation, and services even when contraceptives are provided free of charge 46–48). Occupational engagement also mattered: women engaged in farming were more likely to use FP than unemployed women. This may indicate that farming households perceive FP as a strategy to manage family size in alignment with labor demands. Formal employment was uncommon and not significantly associated with FP use, suggesting that occupation may serve as a proxy for daily activity, household decision-making, and perceived benefits of FP in rural contexts (46,49,50).

Partner education emerged as another key determinant. Women whose partners had secondary education or higher were more likely to use FP, consistent with evidence that educated partners facilitate spousal communication, supportive attitudes, and shared decision-making (33,50,51). Women reporting prior unwanted pregnancies were less likely to use FP, reflecting persistent unmet need and underscoring the importance of early counseling and proactive identification of at-risk women (50,52).

Taken together, these findings indicate that FP utilization in rural Ethiopia is shaped by the interplay of demographic, socioeconomic, geographic, and health system factors. Relatively high FP uptake in the study areas suggests programmatic progress, while persistent inequities including poverty, transport limitations, low partner education, gaps in method awareness, and reliance on home-based sources highlight areas for targeted intervention. HIS may function as a supportive contextual factor, improving data use, monitoring, and follow-up, but further longitudinal or experimental studies are needed to clarify its impact. To promote equitable FP coverage, interventions should integrate community outreach, male involvement, education on the full range of contraceptive options, and strategies to reduce socioeconomic and geographic barriers.

## Strengths and Limitations

### Strengths

This study included a relatively large sample of 840 women of reproductive age, which enhances the reliability and representativeness of the findings. By focusing on Oromia and Gambella, it provides important insights into FP utilization in rural and underserved regions, complementing national survey data. The inclusion of both individual and household-level determinants such as transport access, wealth, occupation, partner’s education, and pregnancy wontedness allowed for a nuanced analysis. The use of multivariable logistic regression strengthened the validity of associations by controlling for confounders. Moreover, linking FP utilization with HIS interventions adds programmatic relevance and highlights the role of data-driven health system strengthening.

### Limitations

As a cross-sectional study, causality between determinants and family planning utilization cannot be inferred. Reliance on self-reported data for FP use and pregnancy intention may have introduced recall bias and social desirability bias. The findings are region-specific and may not be generalizable to urban populations or other Ethiopian regions with different health system contexts. In addition, qualitative perspectives such as women’s perceptions, cultural norms, and provider attitudes were not explored, limiting deeper insight into method preferences and barriers. Finally, unmeasured confounders, including counseling quality, spousal communication, and community influences, may have affected FP uptake but were not directly assessed.

## Conclusion and Recommendation

Family planning utilization in Oromia and Gambella was relatively high (60.9%), with modern methods mainly injectables and implants predominating. However, the continued reliance on short-acting methods highlights gaps in long-term contraceptive planning. FP use was significantly influenced by transport access, household wealth, maternal occupation, partner education, and pregnancy wontedness, underscoring the importance of socioeconomic, geographic, and interpersonal determinants.

Although long-acting method use was higher than national averages, this finding should be interpreted cautiously, as the cross-sectional design does not allow causal attribution to HIS interventions. HIS should therefore be viewed as a supportive system component requiring further study.

In line with Ethiopia’s FP2030 commitments and HIS scale-up efforts, interventions should prioritize community-based outreach and local transport support, strengthen counseling on long-acting methods to promote informed choice, enhance male partner involvement through couple-focused approaches, and use HIS to improve client follow-up and monitoring of unmet need. These targeted actions are essential to advance equitable and sustainable FP utilization in underserved regions.

## Data Availability

The data supporting the findings of this study can be obtained from the corresponding author upon reasonable request

## Abbreviations

AOR: Adjusted Odds Ratio
CBMP: Capacity Building and Mentorship Program
CI: Confidence Interval
DHS: Demographic and Health Survey
DHIS2: District Health Information System 2
FP: Family Planning
HIS: Health Information System
LMICs: Low- and Middle-Income Countries
SSA: Sub-Saharan Africa
WHO: World Health Organization

## Acknowledgements

The authors gratefully acknowledge Jimma University for providing financial and institutional support for this study. We also thank the Oromia and Gambella Regional Health Bureaus, as well as the zonal and district health offices, for their collaboration and facilitation. Our appreciation extends to the data collectors, supervisors, and all study participants for their valuable time and contributions.

## Authors’ contributions

**KH:** Led the study design and conceptualization, coordinated data collection, conducted data analysis and interpretation, and drafted the manuscript. **AW, GT, and MA:** Supported data collection and analysis, contributed to data interpretation, and critically reviewed and revised the manuscript.

## Consent for publication

Not applicable.

